# Toward Precision Medicine in Aortopathies: Genetic Insights from a Brazilian Cohort Through Targeted Next-Generation Sequencing

**DOI:** 10.1101/2023.12.22.23300140

**Authors:** Juliana R. Ferreira, Julia P. Perreira, Anna Paula A. Botelho, Daniele N. Aprijo, Marcelo M. Melo, Helena C. V. Rey, Glauber M Dias

## Abstract

Thoracic aortic diseases (or aortopathies) result from complex interactions between genetic and hemodynamic factors. Often clinically silent, these diseases can lead to lethal complications like aortic dissection or rupture. This study focused on a Brazilian cohort of 79 individuals with thoracic aortic diseases, exploring genetic factors through targeted next-generation sequencing (tNGS) of 15 priority genes. The majority of individuals had non-syndromic aortopathy, with eight diagnosed with Marfan syndrome (MFS). Pathogenic or likely pathogenic variants (PV/LPV) were found in five genes, including FBN1, ACTA2, TGFBR2, MYLK, and SMAD3. Notably, novel variants in FBN1 were identified, contributing to Marfan-like phenotypes. The diagnostic yield for isolated aortopathies was 7.1%, rising to 55.5% for syndromic cases. Variants of uncertain significance (VUS) were identified, emphasizing the need for further research and familial investigations to refine variant classifications. This study provides valuable insights into the genetic landscape of aortopathies in Brazil, aiding early diagnosis and personalized management.

## INTRODUCTION

Aortopathies are diseases that affect the aortic artery and can occur either as an isolated characteristic or as part of a syndromic condition. Their development is linked to a complex interplay of genetic and hemodynamic factors. It is a clinically silent disease with a natural history of lethality, progressing to aortic dissection or rupture, where half of the patients die before reaching emergency services (1,2). The Centers for Disease Control and Prevention (CDC) reports that aortic aneurysms (thoracic and abdominal) are the 15th leading cause of death in individuals over 55 years old and the 19th leading cause of death overall (3). In Brazil, we can observe that the average annual mortality was 12,820 deaths due to aortic diseases, with a total of 128,204 deaths in the last 10 years and a mortality rate of 5.89 (4).

Thoracic aorta diseases (TAD) can be associated with inherited genetic alterations, whereas those affecting the abdominal aorta are more commonly of degenerative origin and related to age and/or risk factors such as hypertension, smoking, dyslipidemia, and gender (5). Hereditary thoracic aortic diseases (HTAD) are characterized by progressive enlargement of the ascending aorta that predisposes to acute dissections (familial thoracic aortic aneurisms /dissections) (6). HTAD can be divided into syndromic and non-syndromic types. Among syndromic TADs, Marfan syndrome (MFS) is the primary type, followed by Loeys-Dietz syndrome (LDS) and Ehlers-Danlos syndrome (EDS), as well as other rarer conditions (7). However, these syndromes contribute to only approximately 5% of all TAD cases, while non-syndromic thoracic aortic diseases (NS-TAD) account for 95% of cases and are further divided into sporadic and familial forms (8).

The genetic factors associated with TADs are inherited as monogenic mutations with an autosomal dominant inheritance pattern (9). Detection of a pathogenic mutation is useful for identifying an individual at risk of a potentially lethal aortic acute event (10). The known genes implicated in this process encode members of the TGFβ signaling pathway (TGFBR1, TGFBR2, TGFB2, SMAD3), components of the aortic wall extracellular matrix’s structure and integrity (FBN1, COL3A1, LOX); or those affecting the function of smooth muscle cells, including actin (ACTA2), myosin (MYH11), myosin light chain kinase (MYLK), and protein kinase cGMP-dependent 1 (PRKG1) (11,12).

The priority genes for TAD can be used to target research and diagnostic panels for aortopathies (13). However, for NS-TADs, the detection rate of variants in these genes is approximately 20-30% (7,10), suggesting the involvement of other mechanisms (genetic or non-genetic) in the pathophysiology. In Brazil, the frequency or incidence of variants in NS-TADs is unknown. We selected a cohort of 79 individuals with isolated and syndromic TADs from Brazil for genetic evaluation through NGS using a targeted gene panel. Here, we report the findings of the genetic screening of a Brazilian cohort of aortopathies.

## SUBJECTS AND METHODS

### Study Population

Patients diagnosed with thoracic aortic aneurysm (TAA), aortic dissection, and aortic coarctation before the age of 60, who were under the care of a quaternary cardiology center in Brazil, were recruited for this study. Inclusion criteria encompassed patients with suspected or confirmed clinical diagnosis of syndromic and non-syndromic aortopathies, all of whom were below 60 years of age (y) at the time of diagnosis. Patients with or without a family history of aortopathies, as well as those clinically suspected of having MFS, LDS and EDS were considered. Exclusion criteria included other causes of aortopathies such as familial dyslipidemia, syphilis, autoimmune and inflammatory diseases, and trauma. A total of 358 patients were evaluated, out of which 103 met the inclusion criteria. After assessing the exclusion criteria, 24 patients were excluded, leaving 79 participants for the study.

### Data Collection

Data collection involved retrospective and prospective evaluation of patients from the date of diagnosis until December 2021. Patients were monitored for the occurrence of aortopathy-related complications (acute aortic syndrome, indication for aortic surgery) and mortality.

### Ethical Approval and Informed Consent

This study received approval from the National Institute of Cardiology Research Ethics Committee (CAAE18868119.5.0000.5272), and all patients provided written informed consent.

### Genetic analysis

Genetic analysis of NS-TAD patients was performed by targeted next-generation sequencing (tNGS) using a panel of 15 priority genes associated with aortic diseases (*ACTA2, COL3A1, FBN1, LOX, MFAP5, MYH11, MYLK, PRKG1, SMAD3, TGFB2, TGFBR1, TGFBR2, EFEMP2, SMAD2, FOXE3*) (13). Sequencing was performed using an Ion Torrent PGM platform (Thermo Fisher Scientific).

Patients with clinical diagnosis (or suspect) of SMF were evaluated by direct sequencing of the coding regions of the FBN1 gene. Reads were compared with the RefSeq NG_008805 using the Geneious Prime Software (Biomatters Ltd).

Detailed information on DNA extraction, targeted next-generation sequencing Ion Torrent PGM (Thermo Fisher Scientific), direct Sanger sequencing are provided in the Supplementary Materials and Methods.

### Variants classification

The variant call files (VCF) were subjected to a series of filters to exclude benign polymorphisms and intronic variants. These filters selected variants located in exons, splice sites, and 3’/5’ UTRs with sequencing coverage ≥ 20x and minor allele frequency (MAF) < 0.0005 in the general population (gnomAD, https://gnomad.broadinstitute.org/). Check of strand bias, indels in homopolymer and high frequency in internal samples were applied to detect and exclude false positives (sequencing artifacts). The remaining variants were classified according to the criteria set forth by the American College of Medical Genetics and Genomics (ACMG) and updates, and the Clinical Genome Resource (ClinGen) (14–17). Sequencing artifact suspects and clinical relevant variants were confirmed by direct sequencing. Reportable variants in this work were pathogenic variants (PV), likely pathogenic variants (LPV), and variants of uncertain significance (VUS).

## RESULTS

We analyzed 79 individuals with TAD. The majority of patients presented a non-syndromic vascular phenotype; eight were diagnosed with MFS. Seventy-two patients were tested by tNGS and 7 MFS individuals were submitted to FBN1 direct sequencing screening. No variants were detected in EFEMP2, FOXE3, SMAD2 and TGFBR1. According to ACMG/AMP guidelines for variant classification we defined 5 PV, 5 LPV, and 19 VUS (Table 1 and 2). Sequencing artifacts and likely benign variants were excluded.

**Table 1.** Pathogenic and likely pathogenic variants.

| ID | Phenotype | Gene | Locus | Transcript | Genotype | Type | Protein impact | Exon | MAF | ACMG Criteria |
| --- | --- | --- | --- | --- | --- | --- | --- | --- | --- | --- |
| AO45<sup>a</sup> | MFS | FBN1 | chr15:48808422 | NM_000138.5 | c.1285C>T | Nonsense | p.Arg429Ter | 11 | NR | **Pathogenic**<br>PVS1, PS4_str<sup>b</sup>, PM2_sup, PP1<sup>c</sup>, PP4 |
| AO56 | Ao dissection | FBN1 | chr15:48760294 | NM_000138.5 | c.4588C>T | Missense | p.Arg1530Cys | 38 | NR | **Likely Pathogenic**<br>PS4_mod<sup>d</sup>, PM2_sup, PP1, PP2, PP3_mod |
| AO71<sup>a</sup> | MFS | FBN1 | chr15:48734026 | NM_000138.5 | c.6055G>T | Nonsense | p.Glu2019Ter | 50 | NR | **Pathogenic**<br>PVS1, PM2_sup, PM6<sup>e</sup>, PP1, PP4 |
| AO84 | Ao dissection | FBN1 | chr15:48800778 | NM_000138.5 | c.1837+1G>C | Sítio de splice | p.? | 15 | NR | **Pathogenic**<br>PVS1_str, PS1<sup>f</sup>, PM2_sup |
| AO85 | Ao dissection | ACTA2 | chr10:90697867 | NM_001613.4 | c.941G>A | Missense | p.Arg314Gln | 8 | NR | **Likely Pathogenic**<br>PP3_str, PM2_sup, PP2 |
| AO86 | Ao dissection/EC | TGFBR2 | chr3:30715721 | NM_001024847.2 | c.1454G>A | Missense | p.Arg485His | 6 | 0.000004 | **Pathogenic**<br>PS4<sup>g</sup>, PM2_sup, PM5, PP3_mod, PP1_str<sup>h</sup> |
| AO87<sup>a</sup> | MFS | FBN1 | chr15:48707855 | NM_000138.5 | c.7929_7930delTG | Del-Frameshift | p.Ser2643ArgfsTer8 | 64 | NR | **Likely Pathogenic**<br>PVS1, PM2_sup |
| AO92 | Ao dissection | MYLK | chr3:123357030 | NM_053025.4 | c.4848delG | Del-Frameshift | p.Ser1617LeufsTer2 | 29 | NR | **Likely Pathogenic**<br>PVS1, PM2_sup |
| AO94<sup>a</sup> | MFS | FBN1 | chr15:48802276 | NM_000138.5 | c.1679G>T | Missense | p.Gly560Val | 14 | NR | **Pathogenic**<br>PS4_sup<sup>i</sup>, PM1, PM2_sup, PP2, PP3_str, PP4 |
| AO99 | Ao aneurysm | SMAD3 | chr15:67477063 | NM_005902.4 | c.872-2A>G | Sítio de splice | p.? | 7 | NR | **Likely Pathogenic**<br>PVS1_str, PS4_sup<sup>j</sup>, PM2_sup |

ACMG, American College of Medical Genetic and Genomic; ID, subject identification; MAF, minor allele frequency in gnomAD; NR, not reported; EC, extracardiac affections; str, strong; mod, moderate; sup, supporting
<sup>a</sup>FBN1 direct sequencing approach
<sup>b</sup>Variant reported in several people with MFS (PMID: 12402346, 18435798, 16835936, 27234404, 19618372)
<sup>c</sup>Variant reported cosegregation with disease in affected family members (PMID: 12402346, 16835936, 18435798)
<sup>d</sup>Variant associated with SMF in 5 non-related people (PMID: 14695540, 17627385, 17663468, 11700157)
<sup>e</sup>Parents are not variant carriers without paternity confirmation
<sup>f</sup>Pathogenic variants with another nucleotide exchange at the same splice site +-1,2 (c.1837+1G>A/T, PMID: 19293843, 15241795)
<sup>g</sup>Variant reported in several people with aortopathies (PMID: 16027248, 16885183, 25644172, 27508510, 23884466, 18852674)
<sup>h</sup>Variant segregates with aortopathies in some families (PMID: 16027248, 16885183)
<sup>i</sup>Variant reported in 2 MFS individuals in ClinVar (https://www.ncbi.nlm.nih.gov/clinvar/variation/803096/)
<sup>j</sup>Variant reported in 1 MFS individuals in ClinVar (https://www.ncbi.nlm.nih.gov/clinvar/variation/1968962/)

The highest incidence of clinically relevant variants was found in the FBN1 gene, with 4 PV and 2 LPV (Figure 1), of these the majority of patients had MFS, and two presented with isolated aortopathy. The others PV/LPV were found once in each of the following genes: ACTA2, TGFB2R, MYLK, and SMAD3. Overall, there were 2 nonsense variants, four missense variants, two frameshift deletions, and two splice site variants. Six variants have been previously reported in association with MFS/TAA or cited in ClinVar database in an aorta disease case. We identified four novel variants in this cohort, of which the truncation variant p.Glu2019Ter in FBN1 was recently reported by our group in association with MFS (18). Hereafter, we detail each variant and the ACMG criteria used for pathogenicity classification.

**Figure 1.**
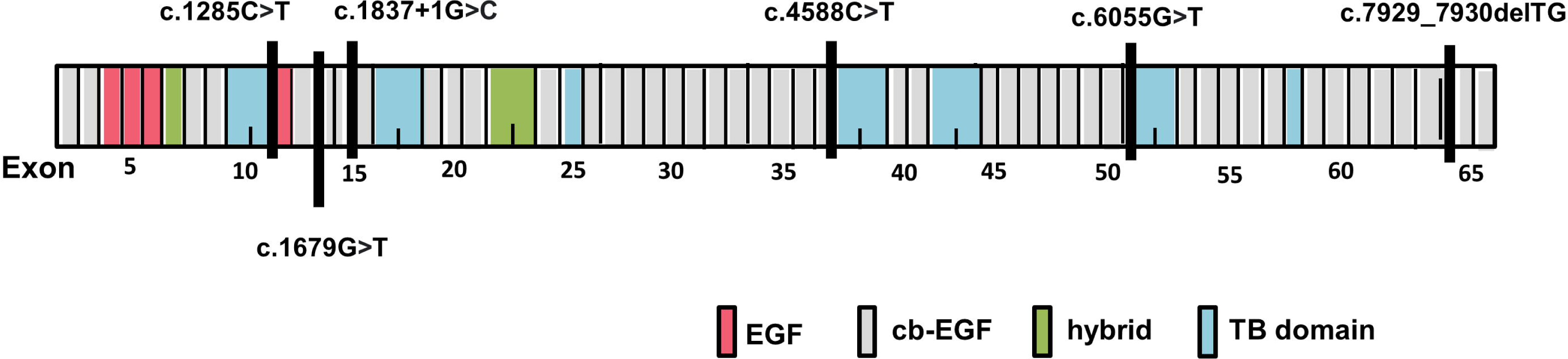
Structure of the FBN1 gene representing the respective domains of the fibrillin-1 protein. The six VP/VPP identified in the FBN1 gene are pointed. EGF: epidermal growth factor-like domains; cb-EGF: EGF-like domains with a calcium-binding domain; TB domain: domains similar to TGF-b binding proteins; hybrid: hybrid containing characteristics of the TB and cb-EGF domains

### FBN1

The missense variants in the FBN1 gene, p.Gly560Val and p.Arg1530Cys, were classified as pathogenic and likely pathogenic, respectively. The FBN1 gene is intolerant to missense variants as demonstrated by the gnomAD Z score value (5.06). Regarding missense variants are a common mechanism of pathogenicity for MFS and TAD (19,20), we applied the PP2 criterion to both variants. REVEL (Rare Variant Ensemble Leaner) score was used for in-silico effect prediction of rare missense variants according to Pejaver et al. (16). The variants p.Gly560Val and p.Arg1530Cys scored 0.945 and 0.871, which fit the PP3_strong and PP3_moderate criteria, respectively. Detection of the p.Gly560Val confirmed the clinical diagnosis of SMF of patient AO94. The p.Arg1530Cys variant have been linked to MFS as well as TAD in various publications (21–29), however, the patient AO56, who carries it, experienced an aortic dissection at the age of 38y without an associated syndromic phenotype.

The nonsense variant p.Arg429Ter in FBN1, identified in a female individual with MFS (AO45), is a known PV. Absent in the gnomAD database, the p.Arg429Ter variant has been reported in several individuals with MFS and has been found to segregate with the disease in families (30–34). This supports the application of PM2_supporting, PS4_Strong and PP1 criteria. The premature termination codon (PTC) generated by the variant is expected to lead to gene dysfunction through protein truncation or nonsense-mediated mRNA decay, which fills the PVS1 criterion.

### TGFBR2

The missense variant c.1454G>A (p.Arg485His) in the Transforming Growth Factor-β receptor gene (TGFBR2) holds well-established clinical significance, being associated with individuals with SMF, LDS, and familial thoracic aortic dissection in multiple studies (35–40). Despite its high prevalence in affected individuals, the frequency of this variant in gnomAD is rare, which supports the application of criteria PS4 and PM2_support, respectively. Additional gnomAD data indicates that the p.Arg485His variant is located in a hotspot region of the TGFBR2 gene, as all 18 VUS/PV described in ClinVar are confined to the region between exon 5 and 8 (PM1). Pannu et al. (36) reported the cosegregation of this variant with the phenotype of familial TAA and dissection with reduced penetrance in two unrelated families (PP1). The REVEL score for the p.Arg485His variant is 0.900, in line with the PP3_moderate criterion. Due to the combined evidence from criteria PS4, PM1, PM2_support, PP1, and PP3_moderate, the p.Arg485His variant is classified as pathogenic.

The variant was detected in a male patient (AO86) diagnosed at 48y with acute aortic syndrome, extracardiac affections (syndrome not diagnosed) and positive family history of cardiac death.

### SMAD3

This gene is part of a group of genes recently associated with HTAD (41). The variant at the canonical splicing site (c.872-2A>G) in the SMAD3 gene results in loss of function of the gene product, which is an established mechanism of pathogenicity in SMAD3 (PVS1) (9,42). This variant is absent from gnomAD (PM2_support) and has a single submission in ClinVar associated with TAA and dissection (PS4_supporting). This PV (PVS1, PM2_supporting and PS4_supporting) was detected in a male patient diagnosed at the age of 35y with a 5.1 cm ascending aortic aneurysm.

### ACTA2

The missense variant p.Arg314Gln (c.941G>A) in the ACTA2 gene has an annotation in ClinVar as a VUS in a TAA case, and is absent in population database (PM2_supporting). The REVEL score of 0.939 indicates the PP3_strong criterion. The ACTA2 gene is intolerant to missense mutations (missense Z score = 3.2), and this type of mutation is a pathogenic mechanism in this gene, which fulfills the PP2 criterion. Due to the combination of PP3_strong, PM2_supporting, and PP2 criteria, the variant has been classified as likely pathogenic. The patient carrying this variant is female and has a positive family history. The diagnosis of TAA occurred at 48y, with an urgent surgical recommendation.

### Novel variants

### FBN1

The FBN1 variant c.1837+1G>C affects the canonical splicing site critical for proper mRNA processing. *In silico* evidence from ASSP (Alternative Splice Site Predictor) and Splice AI indicates the loss of the critical donor site, followed exon skipping and truncated product formation (PVS1). The variant has not been reported in the gnomAD or ClinVar databases, neither in any publication (PM2_support). On the other hand, two different variants at the same locus have been associated with fibrilinopathy/SMF (c.1837+1G>T [43]; c.1837+1G>A [44]). Thus, this variant has been classified as pathogenic based on the combination of PVS1, PM2_support, and PS1 criteria. This variant was detected in a man diagnosed with ascending aortic dissection at 50y and positive family history of cardiac death.

The deletion c.7929_7930delTG in FBN1 alters the reading frame of the mRNA, resulting in a frameshift and generating a PTC, which is expected to lead to the absence of the gene product through nonsense-mediated decay (NMD) (15). This variant has not yet been reported in any publications or databases (PM2_support). Therefore, based on the combination of PVS1 and PM2_support criteria, we classified this variant as likely pathogenic. The patient that harbors this variant is a female with MFS and aortic aneurysm diagnosed at 8y.

### MYLK

A frameshift deletion c.4848delG in the myosin light chain kinase gene (MYLK) has been classified as likely pathogenic based on the following criteria: PVS1 due to gene loss of function through NMD; PM2_support due to absence in the gnomAD database. The patient with this variant is male, with a negative family history. He was diagnosed at 60y with TAA, requiring emergency surgery, and presenting with an ascending aorta diameter of 7.3 cm.

### Diagnosis yield

PV/LPV were identified in five patients out of 70 individuals with non-syndromic aortopathies resulting in an isolated aortopathy diagnosis rate of 7.1% through tNGS. The diagnostic yield for syndromic aortopathies (8 MFS and 1 undefined syndrome) cases was 55.5%, identifying five patients with PV/LPV.

### Variants of Uncertain Significance

Nineteen VUS were detected in this cohort following the classification of reportable variants. Mutations of the following types were observed: missense (11), synonymous (5), one frameshift deletion, and two SNV in UTR. Five variants are novel, and 14 variants have already been reported in ClinVar. The majority of VUS were identified in the FBN1 (7) and MYLK (3) genes, followed by PRKG1 (2), MYH11 (2), and TGFβ2 (2), with one variant in the remaining genes: COL3A1, MFAP5, and LOX (Table 2).

**Table 2.** Variants of uncertain significance.

| ID | Phenotype | Gene | Locus | Transcript | Genotype | Protein impact | Exon | dbSNP | MAF | ACMG Criteria |
| --- | --- | --- | --- | --- | --- | --- | --- | --- | --- | --- |
| AO12 | Ao aneurysm | PRKG1 | chr10:53814273 | NM_006258.4 | c.792C>T | p.Ile264= | 6 | rs1165873275 | 0.000003983 | PM2_sup, BP4 |
| AO32 | Ao aneurysm | TGFB2 | chr1:218519941 | NM_001135599.3 | c.-97delA | p.? | 1 | NR | NR | PM2_sup |
| AO37 | Ao dissection | FBN1 | chr15:48703095 | NM_000138.5 | c.*92T>C | p.? | 66 | rs1038325459 | NR | PM2_sup |
| AO40 | Ao aneurysm | MYH11 | chr16:15815483 | NM_001040114.1 | c.4395C>G | p.Ala1465= | 33 | NR | NR | PM2_sup, BP4, BP7 |
| AO42 | Ao dissection | FBN1 | chr15:48800844 | NM_000138.5 | c.1772A>G | p.Asp591Gly | 15 | rs1380921125 | 0.000003980 | PM1, PM2_sup, PP2, PP3_sup |
| AO44 | Ao aneurysm | MYLK | chr3:123471182 | NM_053025.4 | c.369A>T | p.Val123= | 5 | rs746517311 | 0.000003982 | PM2_sup, BP7 |
| AO45<sup>a</sup> | Ao dissection | FBN1 | chr15:48787450 | NM_000138.5 | c.2547C>G | p.Ile849Met | 22 | rs778258207 | 0.00003581 | PP2, PP4, BP4_mod |
| AO48 | Ao dissection | FBN1 | chr15:48808442 | NM_000138.5 | c.1265G>A | p.Gly422Glu | 11 | rs139968089 | 0.00008139 | PP2 |
| AO55 | Ao dissection | MYH11 | chr16:15810999 | NM_001040114.1 | c.5523C>T | p.Ala1841= | 39 | NR | NR | PM2_sup, BP4, BP7 |
| AO58 | Ao dissection | COL3A1 | chr2:189856420 | NM_000090.4 | c.923G>A | p.Arg308Gln | 13 | rs753589858 | 0.00001592 | PM2_sup, PP2 |
| AO60 | Ao aneurysm | MYLK | chr3:123348391 | NM_053025.4 | c.5044G>A | p.Asp1682Asn | 30 | rs140456660 | 0.00001591 | PM1, PM2_sup |
| AO69 | Ao aneurysm | MFAP5 | chr12:8800764 | NM_003480.4 | c.444delT | p.Arg149GlyfsTer28 | 10 | rs756582294 | 0.00002388 | PVS1_sup |
| AO72 | Ao aneurysm | PRKG1 | chr10:53814271 | NM_006258.4 | c.790A>G | p.Ile264Val | 6 | rs56082459 | 0.00009559 | BS1 |
| AO83 | Ao dissection | FBN1 | chr15:48714264 | NM_000138.5 | c.7455T>G | p.Asp2485Glu | 61 | rs377272529 | 0.00004603 | PM5, PP3_sup |
| AO88 | Ao aneurysm | FBN1 | chr15:48762926 | NM_000138.5 | c.4364T>G | p.Ile1455Ser | 36 | rs397515807 | 0.000003980 | PP3_mod, PS4_sup, PM2_sup, PP2 |
|  |  | LOX | chr5:121412658 | NM_002317.7 | c.670G>C | p.Ala224Pro | 2 | rs1282574280 | NR | PM2_sup |
| AO95 | Ao dissection | MYLK | chr3:123427680 | NM_053025.4 | c.2005T>G | p.Phe669Val | 15 | rs755957947 | 0.00004596 | none |
| AO96 | Ao aneurysm | FBN1 | chr15:48725102 | NM_000138.5 | c.6700G>A | p.Val2234Met | 55 | rs112084407 | 0.0007400 | PP2, BS1, BP4_sup |
| AO99 | Ao aneurysm | TGFB2 | chr1:218607783 | NM_001135599.3 | c.831A>G | p.Arg277= | 5 | rs147960792 | 0.00009245 | none |

ACMG, American College of Medical Genetic and Genomic; ID, subject identification; MAF, minor allele frequency in gnomAD; NR, not reported; EC, extracardiac affections; str, strong; mod, moderate; sup, supporting
<sup>a</sup>FBN1 direct sequencing approach

Two potentially pathogenic VUS in FBN1 were identified in individuals without PV/LPV. These variants did not meet the sufficient criteria for pathogenicity/benignity, due the insufficient available evidences at this time to determine the role of this variation in the disease. However, they stand out from other VUS by nearly reaching the required combination for classification as LPV.

The variant p.Asp591Gly is located in the most missense-intolerant region of the FBN1 gene (O/E missense: 0.41). It has a low population frequency (0.000003980) in gnomAD database, with only one allele detected in the Latin/Amerindian population. We consider PM2_suppoting for MAF<0.002% (45). The REVEL score indicates the PP3_sup criterion. The combination of criteria PM1, PP2, PM2_sup, and PP3_sup for this VUS was insufficient to reach a level of pathogenicity. Patient AO42, who carries this mutation, has been diagnosed with aortic dissection at 48y and has a positive family history related to cardiac death.

The VUS p.Ile1455Ser in the FBN1 gene also had only 1 allele found in gnomAD, but it has already been reported in individuals with MFS/TAD in ClinVar and in a publication (46). The REVEL score was compatible with PP3_mod. The carrier patient (AO88) was primarily diagnosed with ascending aortic aneurysm at the age of 58y.

## DISCUSSION

TAD is a silent and highly morbid condition, and a prompt diagnosis and early intervention can save lives (47,48). Therefore, genetic screening becomes a significant tool for early identification, follow-up, and monitoring of high-risk patients, as well as for determining the course of action (48). In this study, a population with a TAD diagnosis age of ≤60 years was selected, given that this age group exhibits a lower incidence of sporadic TAD (49). An average age of early clinical presentation was observed at 44.6y (±14.3), with a male predominance (63.3%) and 51.9% being of white ethnicity, consistent with observed in other studies (50–52).

In the present study, targeted-Next Generation Sequencing (tNGS) was conducted to identify mutations in genes primarily associated with TAD in 71 patients from a tertiary cardiology hospital. A total of 29 reportable variants were identified, comprising 5 PV, 5 LPV, and 19 VUS. Among isolated aortopathies, our diagnostic yield was 7.1%. Other studies employing tNGS for the investigation of non-syndromic aortopathies reported a diagnostic yield of 4.9% for PV in 1025 patients analyzed (15 sequenced genes) (48) and 16.9% for PV/LPV in 32 genes analyzed (11). Despite the higher yield in the latter work in comparison of our results, SMAD2 was the only additional gene with mutations detected in Thoracic Aortic Aneurysm and Dissection (TAAD) (11).

The above studies have shown that the majority of causal variant were detected in the 11 priority genes. The use of Whole Exome Sequencing (WES) in the investigation for aortopathies does not significantly increase the diagnostic yield of TAD, which generally does not reach 20%. Ziganshin et al. (53) performed WES on 102 TAAD patients and found deleterious variants in 3.9% of cases. Meanwhile, Wolford et al. (52) reported a 10.8% diagnostic yield when conducting WES in cases of aortic dissection or rupture. Yang et al. (54) achieved a diagnostic yield for NS-TAD of 18.7% using an investigative strategy that combined tNGS, MLPA, and WES, with WES adding only 3 individuals to the positive diagnosis group.

Initial clinical suspicion is crucial for guiding genetic testing and achieving better diagnostic yield. In this study, patients with a diagnosis or suspicion of MSF were analyzed through direct sequencing of FBN1, resulting in a diagnostic yield of 50%. This yield is higher than that obtained by Renner et al. (11) with a multigene panel (29.8%) and lower than the 83.5% achieved with the combination of targeted Next-Generation Sequencing (tNGS), Multiplex Ligation-dependent Probe Amplification (MLPA), and Whole Exome Sequencing (WES) (53).

Interpreting genetic sequencing data can be challenging due to variants exhibiting inconsistent characteristics for both pathogenicity and benignity (55). A guideline for interpreting genetic variants in aortopathies has not yet been established. Therefore, efforts dedicated to the careful classification of variants identified in patients with TAD can provide significant contributions to the interpretation and molecular diagnosis of these pathologies within the context of precision medicine (56).

Ten PV/LPV were identified in 5 of the 15 analyzed genes (FBN1, ACTA2, TGFBR2, MYLK, and SMAD3), with four of these variants being reported in association with MFS/TAD for the first time in this study. FBN1 had the highest incidence, with six identified variants, following a similar pattern to other studies (57–59). Mutations in FBN1 were initially associated with MFS (60,61); subsequent studies revealed its association with Marfan-like syndromes (45,57). More recent studies have shown FBN1 variants related to NS-TAD, suggesting a common pathogenesis between NS-TAD and MFS (52,59,62). Cysteine substitutions in the calcium-binding of epidermal growth factor (EGF) like domains represent the majority of pathogenic missense changes associated with MFS (63). However, loss of function (LoF) variants were the principal type of variants discovered in our MFS cohort, followed by a glycine to valine substitution in the EGF-like 8 domain. One missense and one LoF variant were detected in non-syndromic aortopathies. Haploinsufficiency of fibrillin-1 is a pathogenic mechanism for both MFS (64,65) and NS-TAD (54). Missense mutations can also lead to decreased fibrillin secretion due to improper protein folding (66). In these cases, missense mutations and frameshift LoF mutations would share the haploinsufficiency mechanism.

FBN1 was also prevalent in VUS (7). Of all VUS, only one (FBN1-p.Ile849Met) was found in an individual carrying a PV. On the other hand, two missense VUS identified in individuals with isolated aortopathies have the potential to be upgraded to LPV or PV. One of them (FBN1-p.Asp591Gly) is being reported for the first time in this study. The second VUS has previously been reported as an LPV for MFS (46); however, there is still a lack of evidence in the literature to define its burden. Reporting our findings will contribute to potentially elevating a VUS to a causal variant in the future. Investigating families could provide information lacking for the definitive association of VUS with the disease; unfortunately, we did not have access to the families in this study.

In this study, PV/LPV were also found in the genes ACTA2, TGFBR2, MYLK, and SMAD3, for which gene-disease validity is definitive or strong for familial TAAD. Mutations in smooth muscle cell-specific genes (ACTA2, MYLK, MYH11) have been discovered in connection with familial TAAD. This characteristic typically exhibits autosomal dominant inheritance within families, with varying degrees of penetrance and expressivity (67). Actin alpha 2 is a highly conserved protein, and missense variants account for the majority of PV/LPV in this gene. The p.Arg314Gln variant is located in exon 7, the only one where no PV/LPV was annotated.

We identified a novel frameshift variant (c.4848delG) in MYLK with the potential to disrupt protein function and result in loss of allele function through NMD. However, at this time, there is limited evidence to support the haploinsufficiency of this gene, given the limited number of TAAD-associated mutations in MYLK in the literature.

Mutations in the TGFBR2 gene have previously been associated with LDS and TAAD in individuals with Marfanoid features (68,69). Patient AO86 exhibited syndromic features reported as EDS type, but without a conclusive diagnosis. The identified variant has a well-established association with TAD, with most studies reporting its association with isolated aortopathy. However, Law et al. (37) described carriers of the Arg485His variant with features overlapping EDS type 4 (OMIM 130050), such as soft translucent skin, arthralgia, and varicose veins, which could be considered a distinct syndrome.

This investigation provides a comprehensive insight into aortopathies within a Brazilian cohort, highlighting the genetic diversity inherent in these conditions. The implementation of tNGS yielded a notable diagnostic rate, uncovering PV/LPV in genes associated with aortic diseases. The prevalence of mutations in the FBN1 gene, particularly among patients with both syndromic and non-syndromic Marfan-like features, underscores its clinical significance and underscores the necessity for an individualized approach in managing these cases. Furthermore, the identification of novel variants, whether associated with known syndromic phenotypes or not, contributes to the growing understanding of the genetic complexity of aortopathies. These findings underscore the importance of precise genetic screening as a fundamental tool for early intervention and personalized therapeutic strategies, paving the way for significant advancements toward precision medicine in the context of aortopathies.

## Data Availability

All data produced in the present study that are not present in the manuscript are available upon reasonable request to the authors

## Acknowledgments

The authors would like to thank the patients enrolled in this study and the support of INC and FAPERJ.

## Author contributions

**JRF -** Conduction of the work, patient selection and recruitment, clinical evaluation, data collection and analysis, variant analysis, manuscript writing.

**JPP –** Samples processing and NGS.

**APAB -** Samples processing and Sanger sequencing.

**DNA -** Analysis of NGS data and variants interpretation.

**MMM –** Patients selection and recruitment, supervision of clinical evaluation.

**HCVR –** Data analysis, study supervision.

**GMD –** Study idealization, variant analysis, manuscript writing.

## Declaration of interests

The authors declare no competing interests.

## Notes

### Competing Interest Statement

The authors have declared no competing interest.

### Funding Statement

This study was funded by FAPERJ and CNPq

### Author Declarations

Research Ethics Committee of Instituto Nacional de Cardiologia. Eduardo Vera Tibirica

