## Supplementary Materials and Methods for "Toward Precision Medicine in Aortopathies: Genetic Insights from a Brazilian Cohort Through Targeted Next-Generation Sequencing"

**Priority thoracic aortic disease genes screening (prevalence) in a Brazilian adult cohort**

***Genetic analysis***

Genomic DNA was extracted from 4 mL peripheral blood samples using the salting-out method (Longmire et al., 1988). At the conclusion of the process, the eluted DNA in Tris-EDTA buffer was quantified by fluorimetry using the Qubit dsDNA HS Assay kit (Thermo Fisher Scientific) on the Qubit fluorometer.

Genetic analysis of patients with non-syndromic thoracic aortic diseases (DATns) was conducted through targeted Next-Generation Sequencing (tNGS) using a panel of 15 prioritized genes associated with aortic diseases (ACTA2, COL3A1, FBN1, LOX, MFAP5, MYH11, MYLK, PRKG1, SMAD3, TGFB2, TGFBR1, TGFBR2, EFEMP2, SMAD2, FOXE3). Genomic libraries were prepared from 10 ng genomic DNA from each research participant using the Ion AmpliSeq for Chef DL8 kit on the Ion Chef (Thermo Fisher Scientific). Following barcode and adapter ligation, the libraries underwent emulsion PCR using the Ion PGM Hi-Q View Chef 400 kit, loaded onto Ion 318 semiconductor chips, and sequenced on the Ion Torrent PGM platform.

The coding regions of the FBN1 gene in patients clinically diagnosed with Marfan syndrome were directly sequenced using the Sanger method. Additionally, to confirm variants of probable clinical relevance (PV/LPV) and investigate potential artifacts, we also employed the Sanger method for sequencing. Quality control was conducted through electrophoretic analysis of amplified fragments, which were subsequently subjected to sequencing reactions using the BigDye Terminator v3.1 Cycle Sequencing kit (Thermo Fisher Scientific).

Samples were analyzed by capillary electrophoresis on the ABI 3500xl Genetic Analyzer (Thermo Fisher Scientific), and the obtained sequences were compared with the RefSeq NG_008805.2 sequence available on GenBank.
